# Novel Three-Dimensional Imaging Platform for Digital Facial Vitiligo Area Assessment

**DOI:** 10.1101/2025.01.08.25319956

**Authors:** Tiancheng He, Saar Wollach, Sandra L Goss, Mark Loyman, Rowena Bastero, Bethanee J Schlosser, Ming-Chih H Crouthamel, Heidi S Camp, Vardit Eckhouse

**Affiliations:** Data and statistical science, AbbVie Inc, North Chicago, IL, USA; Cherry Imaging Ltd. Yokneam, Israel; Clinical development, AbbVie Inc, North Chicago, IL, USA

## Abstract

**Background:** Vitiligo is a chronic autoinflammatory disorder of the skin due to autoimmune-induced loss of melanocytes in the epidermis that leads to skin depigmentation. Currently, there is no reliable, objective measurement of the depigmented vitiligo area in a clinical trial setting. To assess the extent of facial skin depigmentation in vitiligo patients, subjective scoring methods, e.g., the Facial Vitiligo Area Scoring Index (F-VASI), are used in clinical trials; however, these assessments have low sensitivity and high variability. Here, we developed the novel digital 3D imaging platform for vitiligo lesion quantification and objectively assess facial vitiligo changes.

**Methods:** The 3D imaging platform includes a customized computer visualization app with the handheld stereo optical scanning station. A 3D skin morphology-based image computation model is developed to quantitatively measure facial vitiligo area. To analytically validate this platform, we conducted a synthetic 3D imaging study and a clinical validation study in non-segmental vitiligo patients. In these studies, the 3D imaging platform was used to objectively measure the extent of both synthetic images and facial vitiligo lesions in clinic, and the accuracy and reliability were evaluated.

**Results:** In the synthetic image validation study, 6 synthetic skin models were produced with 4 different synthetic size vitiligo regions and 3 different shades, the average area error is <0.05 cm^2^ for the 6 skin models and <0.04 cm^2^ for the 4 different circles. In the clinical validation study, 25 participants with at least 3 different skin tones (Fitzpatrick scale II-IV) were enrolled. There was high reliability among different scanners and raters, where all intraclass correlation coefficients for intra-scanner reliability, inter-scanner reliability, intra-rater reliability, and inter-rater reliability were > 0.9.

**Conclusions:** This study presented a novel digital 3D imaging platform for facial vitiligo area assessment. The clinical study showed high reliability in the use of non-segmental vitiligo patients. The 3D imaging platform demonstrated high sensitivity in detecting changes in facial vitiligo area over time. The 3D imaging technology can be further developed in interventional clinical trials as an objective and sensitive endpoint to detect changes in facial vitiligo with treatment.

## 1. INTRODUCTION

Vitiligo is a chronic autoinflammatory disorder of the skin characterized by depigmented skin lesions due to autoimmune-induced loss of melanocytes in the epidermis [1]. The disease most commonly presents as white patches of skin with equal impact on men and women [2]. Vitiligo manifests as milky-white patches of pigment loss due to localized melanin destruction. In 2011, an international consensus classified vitiligo into segmental vitiligo (SV) and non-segmental NSV, with the majority of patients (> 90%) exhibiting the non-segmental variant[3,4]. In general, the term vitiligo is being used to describe all forms of NSV, which includes acrofacial, mucosal, generalized, universal, mixed and rare variants. Clinicians often see a distinct border between the normally pigmented areas and the affected depigmented areas [5]. Although the pathogenesis of vitiligo has not yet been clarified, current research can determine many possible factors. In the published literature [6, 7], the main etiologic factors hypothesized by researchers include autoimmune processes, genetic influences, biochemical pathways, and environmental factors. The most well-documented theory is the autoimmune theory, as studies have found clinical associations between vitiligo and autoimmune disorders in several organ systems, including endocrine, gastrointestinal, and neurological disorders. One of the other most important effects of vitiligo is the psychological trauma experienced by people with vitiligo[8]. Vitiligo can greatly contribute to low self-esteem, shame, depression, anxiety, and social isolation, and can also have a profoundly negative impact on quality of life (QoL)[9]. From an economic point of view, vitiligo also imposes a generally heavy financial burden on the patient. Vitiligo has high direct and indirect treatment and care costs, as well as costs associated with other activities such as absenteeism from work. Given these consequences, we advocate that timely diagnosis and treatment of vitiligo is essential.

While efficient medicine and therapy methods are being developed, there is a need to develop a quick and accurate assessment of depigmented skin lesions. So far, various manual scoring methods have been largely developed for efficacy assessment in clinical trial settings. For example, the Vitiligo Area Scoring Index (VASI)[10, 11] and the Vitiligo Extent Score (VES)[12] are two popularly used methods to assess the treatment outcome in the extent of vitiligo. These methods are mainly designed for assessing the whole human body’s depigmentation severity. To specifically assess depigmentation on the face, the facial VASI (F-VASI) score was developed for evaluating treatment efficacy in facial vitiligo[10]. Although this scoring method is useful, it is a still subjective method that is based on the degree of skin depigmentation in those area and is relying on the size of the patient’s fingers or palms to the subject. Previous work also showed its difficulty in assessing certain areas of the face, such as the inside of the eyelids or other small areas of depigmentation and difficulty in assessing pigment with heterogeneous appearance, especially after treatment. Due to these limitations, previous studies also reported the high inter-rater variability and lower reliability when F-VASI was being implemented. An automated digital endpoint for assessing the percentage of facial depigmentation area provides potential solutions to have a more precise and reliable measurement[13]. However, a recent systematic review[14] concluded that validated, fully automated digital imaging technology relies on 2D images for facial vitiligo assessment and highlighted many of the weaknesses without showing full 3D information. In this paper, we described the development of a new 3D digital platform designed to accurately and objectively measure the extent of depigmentation in facial vitiligo areawithin a clinical study setting. The 3D imaging platform includes a customized computer visualization App with the handheld stereo optical scanning station. A 3D skin morphology-based image computation model is developed to automatically measure facial vitiligo area. To analytically validate this platform, we conducted a synthetic 3D imaging study and a clinical validation study in non-segmental vitiligo patients. To validate the system, we tested the accuracy of the platform with the synthetic images and evaluated the consistency and reliability of the 3D digital imaging technology for measuring the areas of facial depigmentation in patients with vitiligo.

## 2. METHODOLOGY

In this study, we developed and validated a novel 3D digital platform for vitiligo area assessments (Cherry Imaging). We also compared 3D measures with multiple synthetic images. This study follows the V3 framework[15], verification, analytical validation, and clinical validation, which were developed by Clinical Trial Transformation Initiatives (CTTI)[16] and the Digital Medicine Society (DiMe)[17] to evaluate the 3D imaging platform which includes the 3D digital stereoscopic scanner (herein 3D scanner) and the imaging model.

### 2.1 Digital Imaging

Briefly, the digital image data in this study are the multispectral high resolution stereoscopic images captured by Cherry Imaging 3D imaging platform [18–21]. The platform contains a 3D scanner, TraceTM software [18], and the Cherry online app for measuring and tracking changes of facial lesions. The 3D scanner uses multi-spectral illuminations to obtain variable characteristics of the skin. The 3D scanner includes an inherent light-emitting diodes (LEDs) with a bandwidth in the order of 10 nm. The capturing process is done by the imaging technician and is similar to a GPS system. The innovative 3D scanner uses stereoscopic technology to lock-on and measure precise 3D coordinates of the targeted area. The TraceTM software creates a gray scaled, low-resolution 3D model which is visualized in real time. Once the real time scan is complete and full face scan is verified, the thousands of 3D high resolution images acquired are combined to create a multi-spectral high resolution (100 microns) 3D reconstruction of the skin’s surface (Figure. 2.1) [22] which is uploaded to the Cherry secured cloud. The high-resolution 3D model is then available on the online Cherry App to allow accurate facial mapping and measurement of different parameters of facial lesions.

**Figure 2.1.**
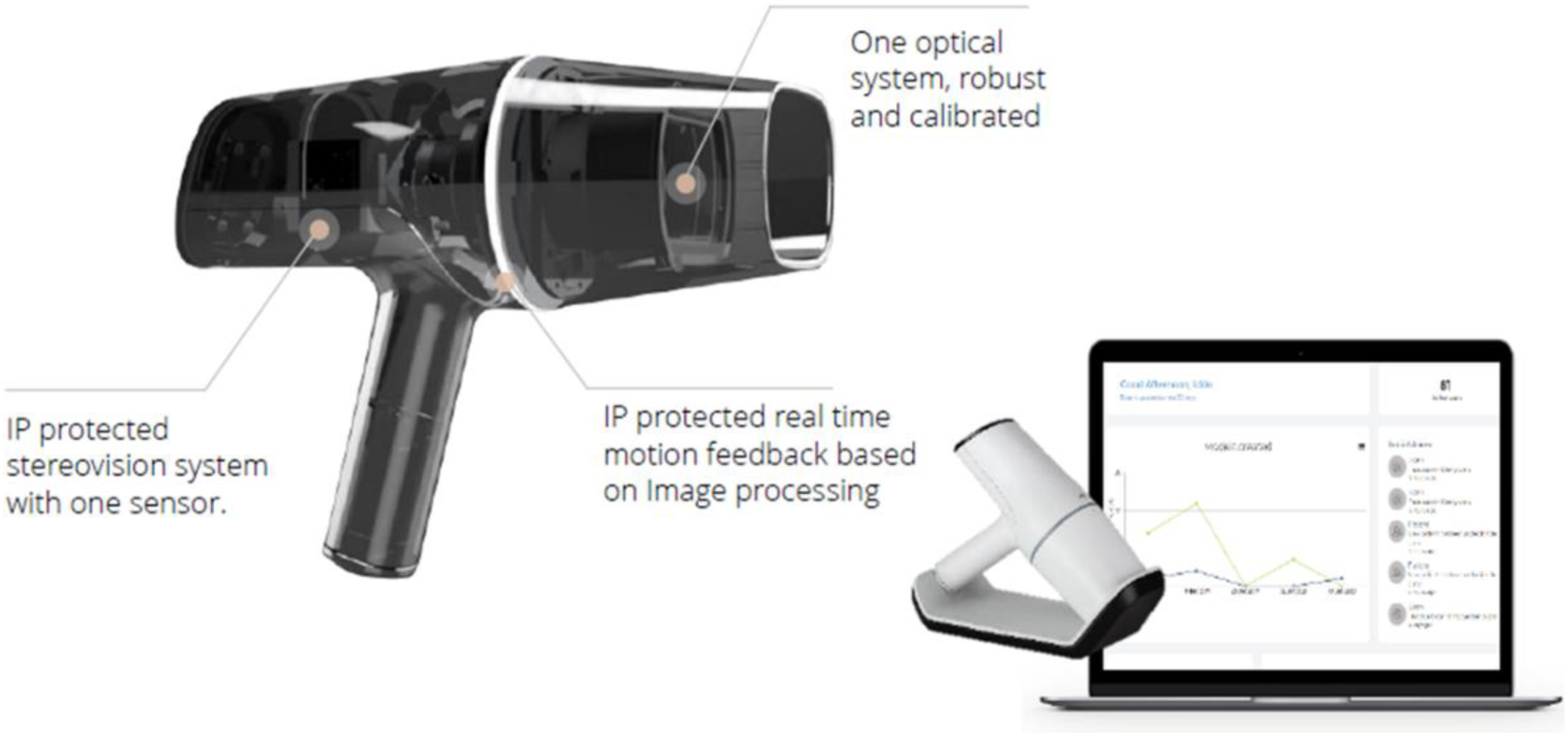
The 3D imaging platform for multi-Layered 3D Reconstruction.

### 2.2 Three-Dimensional Facial Model

In the 3D high resolution color model, the intensity of a 3D pixel is determined by the amount of light for each color (Red, Green, and Blue), that is reflected by the scanned surface. The VTLG color space best distinguishes between normal skin and nonpigmented skin, the color space runs on a scale between 0-1. The degree of VTLG, and therefore pixel intensity, inversely correlates with the degree of pigmentation. That is, skin that is lighter in color (i.e., contains less pigment) reflects more light, resulting in higher pixel intensity values. In contrast, more darkly pigmented skin reflects less light, resulting in lower pixel intensity values (add here something from Marks color model).

### 2.3 Vitiligo Region Measurement

The 3D morphological image analysis method was designed to automate vitiligo region measurement. This method is integrated in the proprietary software by Cherry Imaging in vitiligo subjects to evaluate the reliability using the 3D digital imaging platform. 3D morphological image analysis method is the most commonly applied method of region quantification [23]. This morphological analysis is often performed in conjunction with other methods such as scoring or counting abnormalities. In our study, we present morphological measurements of vitiligo skin depigmentation alongside different types of Fitzpatrick skin tones (FPS) [24]. This was used to thoroughly describe normal skin development and provide a baseline against which to compare abnormal skin depigmentation by vitiligo lesion. Hence, a crucial factor for use of this method is the need for skin lesion segmentation, which was completed in order to quantitatively measure the vitiligo area. This segmentation method has been employed in many studies [19, 21] to remove background and enable measurements of volumes, surfaces areas and distances, so we used this advanced morphological analysis for the required segmentation of complex facial structures. To have a precise skin lesion measurement, this 3D morphological image analysis method was produced by using the threshold-based region-growing tool to select connected regions according to pixel and containing a manual correction to be performed by the physician evaluating the patient.

### 2.4 Image Data Acquisition

The procedure of data acquisition and the determination of vitiligo area using the 3D digital imaging platform contains several steps. The recruited vitiligo patients are assigned a FPS type ranging from 1 to 6. In this study, after the technician scanned, the investigators independently determined and manually selected the vitiligo intensity threshold as shown in Figure 4 for each of the 10 pre-defined anatomic regions of the face while examining the subject. The color space spectrum (VTLG) [25], which is the basis of threshold setting, runs on a scale between 0-1. For the baseline (always selected the first scan), once the high-resolution 3D image is constructed, 10 pre-defined anatomical regions automatically appear on the 3D image. The physician can manually add additional regions of interest and then manually adjusts the intensity threshold for each region on the VTLG scale until all vitiligo lesions in the region are completely and accurately selected. Upon selecting the intensity threshold that optimally distinguishes vitiligo lesions from normal skin for a given subject, the algorithm then combines all 3D pixels with intensities that are greater than the threshold and calculates the total vitiligo area for each region. A composite score of total vitiligo area (cm^2^) on the face is then calculated by summing the automatic vitiligo areas for all regions. In the context of our study, the regions were applied automatically, along with a default threshold value, for all scans. The threshold value was then adjusted according to the observation from physician resulting with the vitiligo measurements (lesion area) (measured in cm^2^) per region.

### 2.5 Clinical Study Design

To validate the system, a clinical validation study was conducted at Herzelia Medical Center, Israel. Twenty-five subjects with various skin tones (Fitzpatrick scale II-IV) participated. The inclusion criteria to enroll included: (1) Healthy subjects, other than vitiligo; (2) Ages 18 and up; and (3) Subjects must read, understand, and sign the Informed Consent. The exclusion criteria were that subjects must not have a history of flash sensitivity or history of epilepsy and/or migraines. The study was reviewed and approved by the institutional review board (IRB) of Herzelia medical center and was conducted in accordance with relevant guidelines and regulations including the Declaration of Helsinki.

All subjects were scanned 3-5 times by three different scanners. The investigator assessed each patient’s Vitiligo lesions using a Woods (UV) lamp. Subjects assessed their own vitiligo condition according to the Patient Global Vitiligo Assessment (PaGVA) score. Each subject’s face was automatically divided into 10 regions (i.e. chin, left eye, right eye, forehead, left lateral cheek, right lateral cheek, left middle cheek, right middle cheek, nose and upper lip). Three raters set the threshold (T) on the model of each patient and the vitiligo area was calculated by the algorithm. Vitiligo area (cm^2^) was calculated for subjects on all scans and all regions. For each patient the 3D first model was uploaded to the cloud, and the 3D model was viewed with the 10 regions in the Vitiligo application. The physician had to set the threshold (0-1) for each one of the regions separately.

## 3. VALIDATION AND RESULTS

To analytically validate digital 3D imaging assessment method, we conducted a synthetic 3D imaging study and a clinical validation study in non-segmental vitiligo patients. In these studies, the 3D imaging platform was used to objectively measure the extent of both synthetic images and facial vitiligo lesions in clinical sites, and accuracy and reliability were evaluated.

### 3.1 Synthetic Image Validation

The objective of the synthetic image study was to determine the level of precision of the Cherry imaging platform by using a predefined size of a color object on a 3D sample imitating skin types between I-VI and different contrasts between the normal skin type color and vitiligo color.

In this synthetic study, the 3D imaging system was tested in Cherry Imaging facilities. As the reference, we used the public dataset of real skin samples [26]. Skin color samples were from an overall of 960 subjects which included 4 ethnic groups (Caucasian, Chinese, Kurdish and Thai) and 4 body locations (forehead, cheek, inner arm, back of hand). Skin color was measured using a spectrophotometer and converted to a device-independent standard color space (The name of color space is CIELAB). For the basic colors to be used, as shown in Figure 3.1, we first estimated the Fitzpatrick skin type using the ITA (individual typology angle) [27] and selected the colors from each FP group to maximize diversity (i.e. FP6: Lowest L* value, FP5: Average, FP4: Lowest b* value, FP3: Highest b* value, FP2: Average, FP1: Highest L* value). Figure 3.2 shows the samples of synthetic images of 6 skin types with vitiligo lesions.

**Figure 3.1.**
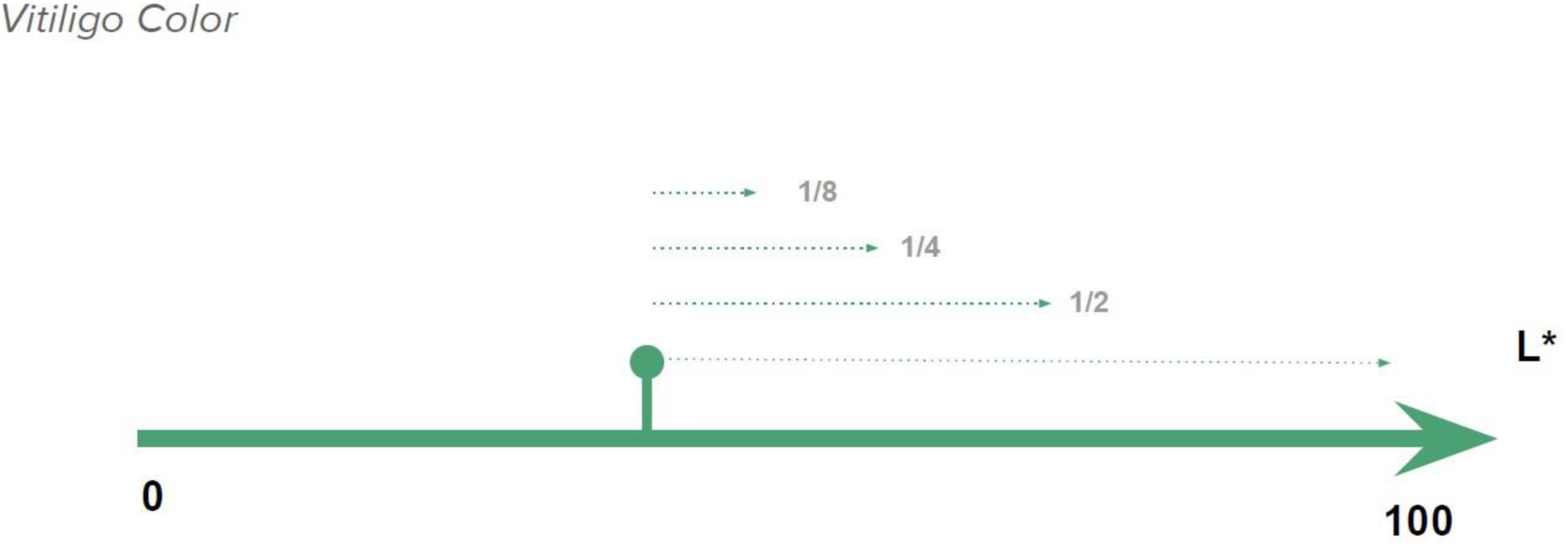
Vitiligo color sampling.

**Figure 3.2.**
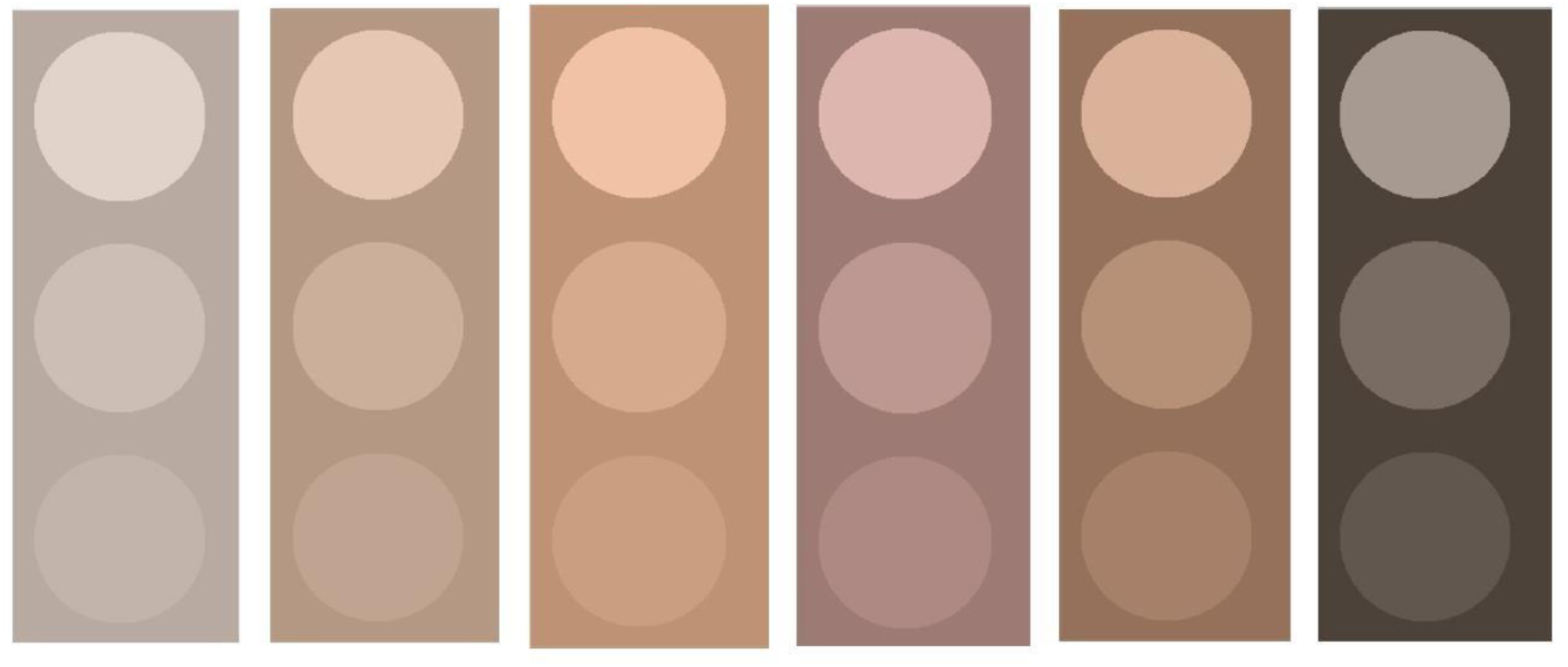
Synthetic images of 6 skin types with vitiligo lesions.

The vitiligo shades are selected by taking the background color (skin types I-VI) and changing the L value (LAB space). In Figure 3.1, the green dot is the background color in LAB space, and the three arrows represent the three different contrast shades. The L is defined in the range between 0 – 1.

Six synthetic models were produced with 4 different synthetic size vitiligo regions and 3 different shades, imitating skin types I-VI. Each model had circles of the following diameters: 0.5cm, 1 cm, 2cm and 4cm. An example of skin types I and VI are presented in Figure 3.3. Supplementary Table 1 illustrates all the shape information about the synthetic models.

**Figure 3.3.**
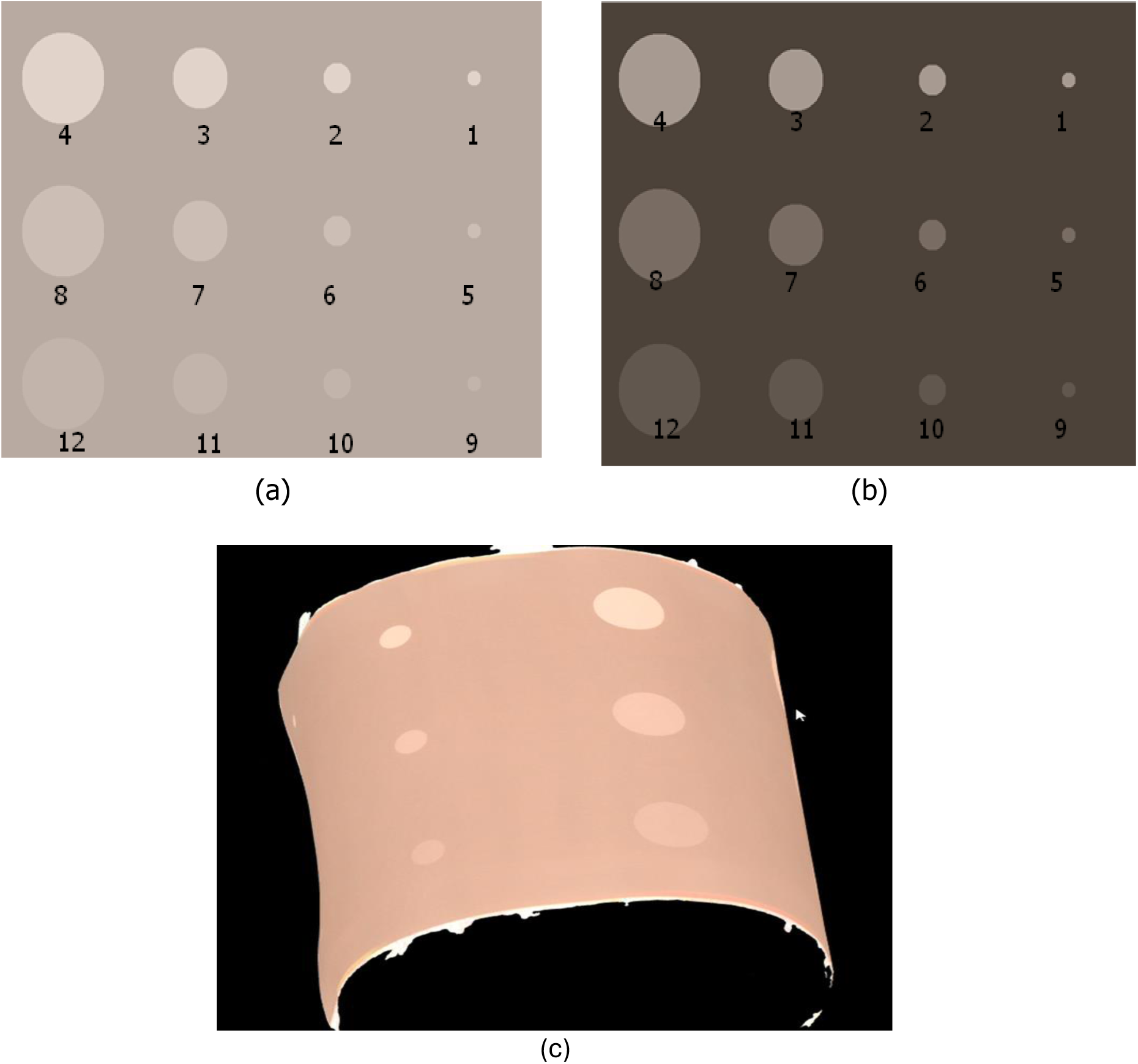
An example of synthetic images with different skin types, (a) skin type I, (b) skin type VI, (c) a curved scanned surface.

To generate the synthetic data using 3D imaging system, each skin type model was scanned 1 time, the model of skin type III was scanned another time while set on a curved surface as shown in Figure 3.3(c). The threshold was set for each of the 12 regions separately and the algorithm measured the vitiligo area for each one of the regions. For each circle the threshold was set separately, and the area was calculated accordingly. The correlation between the real area of the circles (measured by a caliber) and the calculated area for the different skin types was determined. We measured the average error between the real area and the calculated area for the 6 different skin types. The results are presented in Table 3.1. Table 3.2 summarized the results for average error for the 4 different circles. The average error was calculated as a function of the contrast. We divided the contrast into 4 regions, 0-5, 5-10, 10-20 and 20-25, the results are presented in Table 3.3. We also validated the effect of threshold on area measurements. Table 3.4 summarizes the accuracies of the models with fixed threshold for all 3 contrasts (circle 4, 8, and 12) for skin types I and VI. In conclusion, the results of synthetic data show that the 3D image system can reach a promising accuracy and reliability in terms of the measurement errors and correlations between system measured area and the real area.

**Table 3.1.** Correlation and average error for the 6 different skin types.

| **Skin Type** | **Correlation** | **Average error** |
| --- | --- | --- |
| I | 0.999 | 0.086 |
| II | 0.999 | 0.050 |
| III | 0.999 | 0.036 |
| III<br>Curved | 0.999 | 0.109 |
| IV | 0.999 | 0.021 |
| V | 0.999 | 0.015 |
| VI | 0.999 | 0.020 |

**Table 3.2.** Average error as a function of the circle sizes.

| **Area** | **Average Error [cm<sup>2</sup>]** |
| --- | --- |
| 0.19 | 0.011 |
| 0.78 | 0.019 |
| 3.14 | 0.037 |
| 12.56 | 0.126 |

**Table 3.3.** Average error as a function of the contrast (we divided the contrast range into 4 regions)

| **Contrast** | **Average Error [cm<sup>2</sup>]** |
| --- | --- |
| 0 to 5 | 0.105 |
| 5 to 10 | 0.037 |
| 10 to 20 | 0.041 |
| 20 to 25 | 0.013 |

**Table 3.4.**
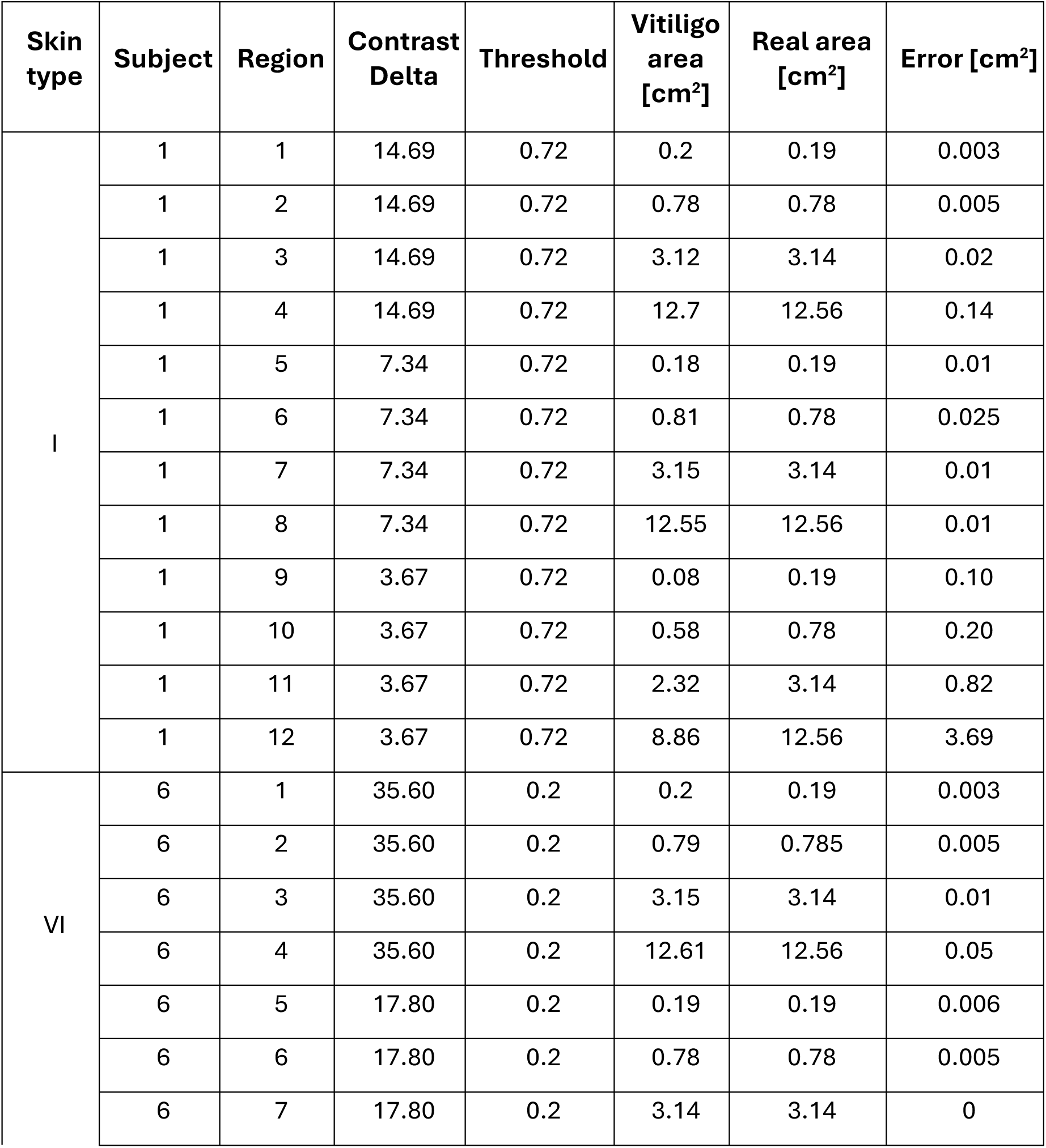
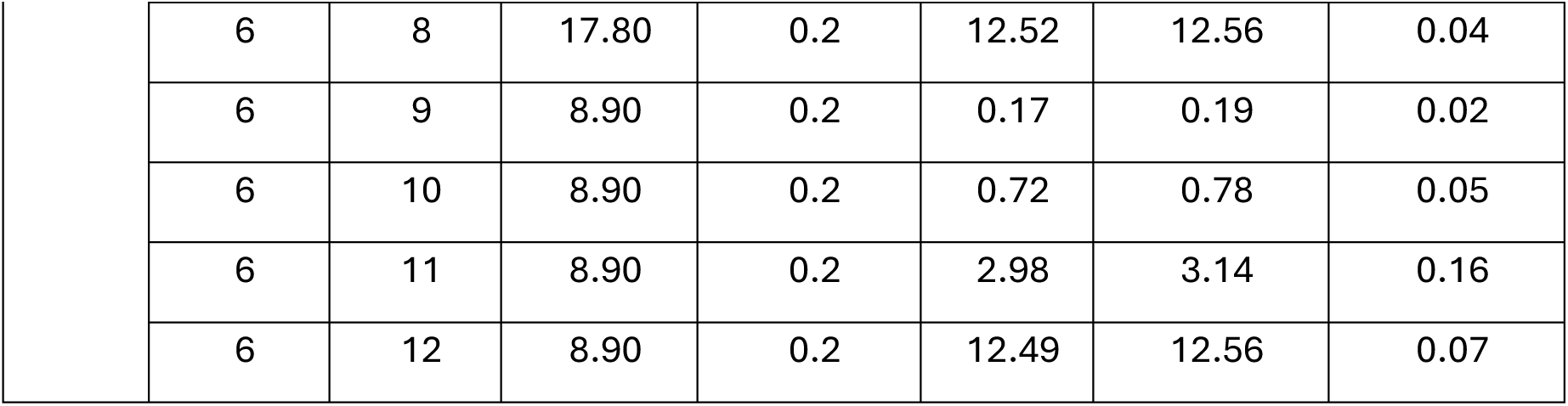
Fixed threshold for all 3 contrasts (circle 4, 8, and 12) for skin types I and VI.

### 3.2 Clinical Validation

To validate the 3D imaging platform in clinical validation study, we scanned patients with Vitiligo lesions on the face to evaluate the precision, variability, and the dynamic range of face Vitiligo lesion To evaluate the performance, we determined the intra-rater reliability and inter-rater variability of the vitiligo measurements and determined the dynamic range detection (sensitivity) in various skin tones.

#### 3.2.1 Reliability Analyses on Clinical Validation Study Data

##### I. Intra-Rater Reliability

To assess the intra-rater reliability in vitiligo area measurement, in each of the 10 regions, across the three measurements varying in threshold, we employed the Intraclass Correlation Coefficient (ICC). ICC estimates were calculated using SPSS statistical package version 24 (SPSS Inc, Chicago, IL) based on a single measurement, absolute-agreement, 2-way mixed-effects model, following Shrout and Fleiss [28], and Koo & Lee [29]. Table 3.5 presents all single measurement correlations in detail. As can be observed from the table, almost all coefficients were above 0.9, pointing at excellent reliability.

**Table 3.5.** Intra-Rater Reliability Represented by Intraclass Correlation Coefficients.

|  | Complete set of subjects (N=9) |  |
| --- | --- | --- |
|  | Full Data | No Vitiligo area =0 measurements |
| Region 1 | ** | ** |
| Region 2 | .989 [.966-.997] | .989 [.966-.997] |
| Region 3 | .995 [.986-.999] | .982 [.930-.997] |
| Region 4 | .983 [.948-.996] | .972 [.905-.995] |
| Region 5 | .984 [.954-.996] | .924 [.579-.998] |
| Region 6 | .988 [.966-.997] | ** |
| Region 7 | .999 [.998-1.000] | .999 [.988-1.000] |
| Region 8 | .999 [.996-1.000] | .997 [.973-1.000] |
| Region 9 | .937 [.737-.993] | .937 [.737-.993] |
| Region 10 | .997 [.991-.999] | .981 [.677-1.000] |

*Note.* Values in brackets represent the 95% confidence interval; \*Correlation coefficient was not computed due to zero variance items; \*\*Too few cases for analysis ( $N < 2$ )

##### II. Inter-Rater Reliability

To assess the inter-rater reliability in vitiligo area measurement, in each of the 12 regions, across the three raters, we employed the Intraclass Correlation Coefficient (ICC). Table 3.6 presents all single measurement correlations in detail. As can be observed from the table, all coefficients were above 0.9, pointing to excellent reliability.

**Table 3.6.** Inter-Rater Reliability Represented by Intraclass Correlation Coefficients.

| | Complete set of subjects ($N=25$) | |
| --- | --- | --- |
|  | Full Data | No Vitiligo area =0 measurements |
| Region 1 | .997 [.993-.999] | .999 [.990-.999] |
| Region 2 | .944 [.894-.974] | .944 [.894-.974] |
| Region 3 | .959 [.922-.981] | .934 [.866-.972] |
| Region 4 | .998 [.995-.999] | .997 [.993-.999] |
| Region 5 | .995 [.990-.998] | .992 [.979-.998] |
| Region 6 | .997 [.994-.999] | .996 [.985-.999] |
| Region 7 | .999 [.999-1.000] | .999 [.996-1.000] |
| Region 8 | .997 [.994-.998] | .987 [.960-.997] |
| Region 9 | .981 [.959-.992] | .976 [.945-.991] |
| Region 10 | .993 [.986-.997] | .956 [.821-.995] |

##### III. Intra-Scanner Reliability

To assess the intra-scanner reliability in vitiligo area measurement, of each of the four scanners, in each of 10 regions, across the five models, we employed the Intraclass Correlation Coefficient (ICC). Noteworthy, scanner 1 scanned subjects 1 through 5 and 10 through 25, and scanner 4 scanned subjects 6 through 9, while scanners 2 and 3 scanned the complete set of subjects. Table 3.7 presents all single measurement correlations in detail on all measurements, and Table 3.8 presents all single measurement correlations in detail on all measurements excluding vitiligo area = 0. As can be observed from the tables, almost all coefficients were above 0.9, pointing at excellent reliability, with the exclusion of region 6 – right lateral cheek, scanner 3, when not including measurements of no vitiligo, among all subjects (0.89).

**Table 3.7.** Intra-Scanner Reliability Represented by Intraclass Correlation Coefficients – Complete Set of Subjects (N=25), Full Data.

|  | Scanner 1 | Scanner 2 | Scanner 3 | Scanner 4 |
| --- | --- | --- | --- | --- |
| Region 1 | .992 [.982-.997] | .994 [.987-.998] | .993 [.985-.997] | * |
| Region 2 | .992 [.982-.997] | .993 [.985-.997] | .992 [.982-.997] | .985 [.894-1.000] |
| Region 3 | .993 [.985-.998] | .996 [.991-.998] | .996 [.992-.999] | .974 [.763-1.000] |
| Region 4 | .997 [.992-.999] | .997 [.994-.999] | .998 [.996-.999] | * |
| Region 5 | .993 [.986-.998] | .991 [.980-.997] | .994 [.987-.998] | * |
| Region 6 | .974 [.943-.991] | .956 [.909-.983] | .920 [.841-.970] | .968 [.793-1.000] |
| Region 7 | .996 [.991-.999] | .998 [.996-.999] | .996 [.991-.999] | * |
| Region 8 | .999 [.997-.999] | .996 [.992-.999] | .992 [.984-.997] | .999 [.989-1.000] |
| Region 9 | .993 [.985-.998] | .994 [.988-.998] | .994 [.987-.998] | .980 [.853-1.000] |
| Region 10 | .998 [.995-.999] | .996 [.991-.998] | .995 [.989-.998] | 1.000 [.999-1.000] |

*Note.* Values in brackets represent the 95% confidence interval; \*Correlation coefficient was not computed due to zero variance items

**Table 3.8.** Intra-Scanner Reliability Represented by Intraclass Correlation Coefficients - Complete Set of Subjects (N=25), no Vitiligo area = 0 Measurements.

|  | Scanner 1 | Scanner 2 | Scanner 3 | Scanner 4 |
| --- | --- | --- | --- | --- |
| Region 1 | .987 [.968-.997] | .990 [.974-.997] | .989 [.973-.997] | ** |
| Region 2 | .990 [.977-.997] | .991 [.981-.997] | .992 [.982-.997] | .985 [.894-1.000] |
| Region 3 | .985 [.959-.996] | .991 [.978-.998] | .994 [.984-.998] | .974 [.763-1.000] |
| Region 4 | .996 [.990-.999] | .997 [.992-.999] | .998 [.994-.999] | ** |
| Region 5 | .989 [.955-.999] | .981 [.926-.999] | .967 [.849-.999] | ** |
| Region 6 | .933 [.718-.998] | .928 [.779-.991] | .886 [.674-.985] | ** |
| Region 7 | .995 [.965-1.000] | .998 [.983-1.000] | .995 [.959-1.000] | ** |
| Region 8 | .997 [.983-1.000] | .988 [.951-.999] | .977 [.910-.998] | ** |
| Region 9 | .991 [.979-.997] | .992 [.983-.997] | .992 [.983-.997] | .980 [.853-1.000] |
| Region 10 | .994 [.984-.999] | .991 [.976-.998] | .987 [.968-.997] | ** |

*Note.* Values in brackets represent the 95% confidence interval; \*\*Too few cases for analysis (N<2)

##### IV. Inter-Scanner Reliability

Following the high Intra-Scanner reliabilities, we proceeded to compute average scores across models, for each region, each scanner and each participant. To assess the **inter-scanner reliability** in vitiligo area measurement, in each of 10 regions, across three scanners (scanners 1 through 3 for participants 1-5 and 10-25, and scanners 2 through 4 for participants 6-9), we employed the Intraclass Correlation Coefficient (ICC). Noteworthy, for scanner 4, who scanned only 4 subjects, as noted before, in many of the cases the measurement was zero, so when examining the reliability of scanners 2 through 4 excluding zero measurements of Vitiligo, there were mostly too few cases left for analysis (in fact, only in two of the 10 regions scanner 4 had non-zero data for all 4 subjects). Therefore, in the report on reliabilities with no Vitiligo = 0, we include only scanners 1 through 3. Table 3.9 present all single measurement correlations in detail. As can be observed from the tables, almost all coefficients were above 0.9, pointing at excellent reliability, with the exclusion of region 8 – right middle cheek, scanners 2-4, when not including measurements of no vitiligo, among all subjects (.56), and region 9 – nose, scanners 2-4, when not including measurements of no vitiligo, among skin type III-IV subjects (.36).

**Table 3.9.** Inter-Scanner Reliability Represented by Intraclass Correlation Coefficients - Complete Set of Subjects (N=25)

|  | Full Data |  | No Vitiligo area =0 measurements |
| --- | --- | --- | --- |
|  | Scanners 1-3 | Scanners 2-4 | Scanners 1-3 |
| Region 1 | .998 [.996-.999] | .999 [.996-1.000] | .998 [.995-.999] |
| Region 2 | .993 [.978-.997] | .983 [.908-.999] | .992 [.974-.997] |
| Region 3 | .996 [.990-.998] | .992 [.948-.999] | .994 [.983-.998] |
| Region 4 | .999 [.998-.999] | .999 [.996-1.000] | .999 [.997-.999] |
| Region 5 | .998 [.996-.999] | 1.000 [.999-1.000] | .997 [.991-.999] |
| Region 6 | .999 [.998-1.000] | 1.000 [.999-1.000] | .999 [.995-1.000] |
| Region 7 | .997 [.994-.999] | .999 [.996-1.000] | .993 [.972-.999] |
| Region 8 | .998 [.997-.999] | .997 [.983-1.000] | .996 [.987-.999] |
| Region 9 | .998 [.996-.999] | .964 [.783-.998] | .998 [.995-.999] |
| Region 10 | .997 [.994-.999] | .999 [.992-1.000] | .993 [.983-.998] |

*Note.* Values in brackets represent the 95% confidence interval; \*Correlation coefficient was not computed due to zero variance items; \*\*Too few cases for analysis (N<2)

## 4. DISCUSSION

In this paper, we validated the novel digital 3D imaging method for vitiligo area assessment by using the new 3D imaging platform. The clinical study showed high reliability in the 3D imaging modality determining facial vitiligo area. The 3D imaging platform also demonstrated high accuracy and sensitivity in detecting vitiligo area in different skin types over time by validating the measurements in various synthetic data.

Different scales have been proposed in medicine for assessing the extent and severity of vitiligo, but of course each scale has its own strengths and limitations. All these techniques require the experience of the medical professionals combined with a visual assessment in order to make quantitative measurements. Medical professionals often need to assess the size, location, and progression of depigmented patches on the skin. For current vitiligo treatment assessments, the quantitative scores derived from these assessments play a crucial role in tracking the progression of the disease and evaluating the effectiveness of therapeutic interventions. One popular method is the Facial Vitiligo Area Scoring Index (F-VASI), which takes the facial skin affected by vitiligo and scores it based on the percentage of overall facial skin. However, this technique does not provide a comprehensive assessment of irregularly shaped patches or patches located in unnoticeable areas and is highly subjective. Another index commonly used in clinical settings is the Vitiligo Extent Score (VES). The VES uses stenciled photographs to assess vitiligo on all parts of the body, but does not include the back of the scalp, the soles of the feet, or the palms of the hands [30]. Although the VES has a template as a specific quantitative criterion, it is also subjective in the same way as the F-VASI. Therefore, the biggest limitation of the current clinical scales is the high potential for variability in assessing vitiligo.

In addition to current clinical scales, imaging techniques play an effective role in assessing structural changes in the epidermis of vitiligo. Since imaging techniques can provide very detailed images with more accurate and objective information about the distribution and extent of depigmentation, this is particularly important for detecting the progression of vitiligo and monitoring the efficacy of treatments as well as deepening the understanding of the underlying pathogenesis. Currently there are many effective imaging techniques for the detection of vitiligo, such as Ultraviolet light photography[31], Optical coherence tomography[32], Visible light and digital photography[33], but the main limitations of these imaging techniques are the requirement of a specific operating environment and the cost of acquiring imaging equipment can be, as well as the need for experienced professionals to follow rigorous procedures that cannot be performed in any healthcare setting. When utilizing the results of these imaging techniques, researchers are using computer-aided imaging analysis, specifically training machine learning algorithms [34, 35] to analyze skin images and provide highly accurate assessments of the extent and progression of the vitiligo. Unlike traditional imaging techniques that rely on manual interpretation, machine learning algorithms can provide a consistent and unbiased assessment of the skin. This is particularly useful for large-scale studies and for tracking skin changes over time. However, the main limitation to using machine learning techniques to diagnose vitiligo is due to their high dependence on training data [36]. The ability to use a portable imaging device with proven image processing techniques in the field of diagnosing vitiligo would have many advantages.

This is the first clinical study to utilize portable 3D imaging to assess vitiligo in a quantitative and objective trial. However, we also identified some limitations in the results of the current study. First, this is a non-interventional clinical validation study with a limited sample size of vitiligo patients. To implement this technology in clinical use, a follow-up study utilizing portable 3D imaging for clinical applicability of treatment for patients with vitiligo at different treatment plans is needed. Second, there was a relative imbalance in the patient cohort with different skin tones, a common challenge for validation all vitiligo detection techniques. Compared with the vitiligo imaging detection techniques that have been proposed so far, our 3D imaging technique may have better accessibility because of its portability and low requirement of clinical environment and may be more suitable for wide use and clinical dissemination. More research is needed to assess of the reliability of this 3D imaging technique across skin types and in multiple clinics.

## 5. CONCLUSION

This study showed high accuracy and reliability in the measurement of facial vitiligo area determined by a novel 3D imaging modality. The digital 3D platform demonstrated greater sensitivity for detecting change in facial vitiligo area over time in the clinical validation study. It enables objective measurement of facial vitiligo area and provides visual confirmation of vitiligo changes. Further evaluation of the system is required in more patients and across all Fitzpatrick skin phototypes.

## Data Availability

All data produced in the present work are contained in the manuscript

## APPENDIX

**Supplementary Table 1.** summarized information about the RGB values for the 6 samples along with the contrast.

| Skin type | Item | R | G | B | Contrast |
| --- | --- | --- | --- | --- | --- |
| 6 | Background | 76.71456 | 65.96169 | 57.69942 | 0 |
|  | Circle1-4 | 166.8062 | 153.8977 | 144.3305 | 35.6 |
|  | Circle 5-8 | 120.2919 | 108.3283 | 99.34292 | 17.8 |
|  | Circle 9-12 | 98.0995 | 86.69799 | 78.05166 | 8.9 |
| 5 | Background | 149.4343 | 112.3828 | 89.85808 | 0 |
|  | Circle1-4 | 218.353 | 176.9673 | 152.505 | 24.75 |
|  | Circle 5-8 | 183.3834 | 144.0234 | 120.4727 | 12.375 |
|  | Circle 9-12 | 166.2764 | 128.0302 | 104.9763 | 6.1875 |
| 4 | Background | 157.4384 | 122.2347 | 115.9023 | 0 |
|  | Circle1-4 | 220.5089 | 182.2201 | 175.2565 | 22.75 |
|  | Circle 5-8 | 188.5265 | 151.695 | 145.0252 | 11.375 |
|  | Circle 9-12 | 172.8662 | 136.8246 | 130.3172 | 5.6875 |
| 3 | Background | 188.543 | 145.6637 | 117.2714 | 0 |
|  | Circle1-4 | 240.5084 | 194.4003 | 164.4797 | 18.12 |
|  | Circle 5-8 | 214.2754 | 169.7127 | 140.5279 | 9.06 |
|  | Circle 9-12 | 201.345 | 157.6053 | 128.809 | 4.53 |
| 2 | Background | 179.6491 | 151.6274 | 132.2229 | 0 |
|  | Circle1-4 | 229.2345 | 199.3248 | 178.9233 | 17.65 |
|  | Circle 5-8 | 204.1818 | 175.1762 | 155.2548 | 8.825 |
|  | Circle 9-12 | 191.8486 | 163.324 | 143.656 | 4.4125 |
| 1 | Background | 183.8085 | 170.3922 | 160.9936 | 0 |
|  | Circle1-4 | 224.737 | 210.6951 | 200.9197 | 14.69 |
|  | Circle 5-8 | 204.0842 | 190.3449 | 180.7522 | 7.345 |
|  | Circle 9-12 | 193.8979 | 180.3174 | 170.8202 | 3.6725 |

**Supplementary Table 2.** Intra-Rater Reliability Represented by Intraclass Correlation Coefficients according to Skin Types.

| Skin Type I-II subjects (N=5) |  | Skin Type III-IV subjects (N=4) |  |
| --- | --- | --- | --- |
| Full Data | No Vitiligo area =0 measurements | Full Data | No Vitiligo area =0 measurements |
| .976 [.747-1.000] | ** |  |  |
| .992 [.963-.999] | .992 [.963-.999] | .989 [.937-.999] | .989 [.937-.999] |
| .995 [.974-.999] | .987 [.921-.999] | .997 [.983-1.000] | .974 [.711-1.000] |
| .957 [.808-.995] | .905 [.431-.997] | .997 [.979-1.000] | .997 [.979-1.000] |
| * | ** | .972 [.859-.998] | .924 [.579-.998] |
| * | ** | .988 [.939-.999] | ** |
| .987 [.943-.999] | ** | .999 [.996-1.000] | ** |
| .999 [.996-1.000] | .999 [.986-1.000] | .999 [.991-1.000] | ** |
| .947 [.730-.996] | .947 [.730-.996] | ** | ** |
| * | ** | .996 [.963-1.000] | .981 [.677-1.000] |
| * | ** | .975 [.836-.999] | .800 [.080-1.000] |
| .951 [.796-.994] | .891 [.530-.992] | .999 [.996-1.000] | ** |

*Note.* Values in brackets represent the 95% confidence interval; \*Correlation coefficient was not computed due to zero variance items; \*\*Too few cases for analysis (N<2)

**Supplementary Table 3.** Inter-Rater Reliability Represented by Intraclass Correlation Coefficients according to Skin Types.

| Skin Type I-II subjects (N=8) |  | Skin Type III-IV subjects (N=17) |  |
| --- | --- | --- | --- |
| Full Data | No Vitiligo area =0 measurements | Full Data | No Vitiligo area =0 measurements |
| .978 [.887-.998] | .970 [.804-.999] | .998 [.992-.999] | .997 [.987-1.000] |
| .901 [.710-.977] | .901 [.710-.977] | .952 [.891-.982] | .952 [.891-.982] |
| .986 [.954-.997] | .976 [.910-.996] | .950 [.891-.981] | .920 [.813-.972] |
| .979 [.933-.995] | .959 [.803-.997] | .998 [.995-.999] | .997 [.993-.999] |
| .906 [.735-.978] | ** | .996 [.991-.999] | .995 [.985-.999] |
| * | ** | .997 [.992-.999] | .996 [.985-.999] |
| .999 [.997-1.000] | ** | .999 [.998-1.000] | .998 [.995-1.000] |
| .998 [.992-.999] | .950 [.795-.994] | .996 [.992-.999] | .974 [.912-.995] |
| .967 [.890-.994] | .950 [.795-.994] | .981 [.950-.994] | .978 [.941-.993] |
| * | ** | .992 [.981-.997] | .956 [.821-.995] |
| * | ** | .999 [.997-1.000] | .998 [.992-1.000] |
| .980 [.937-.996] | .945 [.767-.994] | .992 [.982-.997] | .986 [.962-.996] |

*Note.* Values in brackets represent the 95% confidence interval; \*Correlation coefficient was not computed due to zero variance items; \*\*Too few cases for analysis (N<2)

**Supplementary Table 4.** Intra-Scanner Reliability Represented by Intraclass Correlation Coefficients - Skin Type I-II subjects (N=8), Full Data.

|  | Scanner 1 | Scanner 2 | Scanner 3 |
| --- | --- | --- | --- |
| Region 1 | .993 [.976-.999] | .997 [.989-.999] | .994 [.983-.999] |
| Region 2 | .976 [.920-.997] | .978 [.932-.996] | .987 [.962-.997] |
| Region 3 | .987 [.956-.998] | .991 [.972-.999] | .994 [.982-.999] |
| Region 4 | .998 [.988-1.000] | .996 [.985-.999] | .992 [.977-.998] |
| Region 5 | * | * | * |
| Region 6 | .967 [.891-.996] | .951 [.858-.992] | .915 [.782-.982] |
| Region 7 | * | * | * |
| Region 8 | .972 [.907-.997] | .991 [.972-.999] | .981 [.945-.996] |
| Region 9 | .988 [.959-.999] | .988 [.961-.998] | .985 [.957-.997] |
| Region 10 | .997 [.990-1.000] | .994 [.981-.999] | .996 [.988-.999] |

*Note.* Values in brackets represent the 95% confidence interval; \*Correlation coefficient was not computed due to zero variance items

**Supplementary Table 5.** Intra-Scanner Reliability Represented by Intraclass Correlation Coefficients - Skin Type III-IV subjects (N=17), Full Data.

|  | **Scanner 1** | **Scanner 2** | **Scanner 3** |
| --- | --- | --- | --- |
| Region 1 | .987 [.965-.997] | .986 [.963-.997] | .989 [.970-.998] |
| Region 2 | .994 [.984-.999] | .997 [.993-.999] | .995 [.985-.999] |
| Region 3 | .997 [.991-.999] | .999 [.997-1.000] | .998 [.995-1.000] |
| Region 4 | .997 [.991-.999] | .997 [.993-.999] | .998 [.995-1.000] |
| Region 5 | .993 [.980-.998] | .989 [.971-.998] | .992 [.976-.999] |
| Region 6 | .995 [.987-.999] | .977 [.937-.995] | .975 [.929-.995] |
| Region 7 | .996 [.989-.999] | .998 [.995-1.000] | .996 [.988-.999] |
| Region 8 | .999 [.997-1.000] | 1.000 [.999-1.000] | .999 [.998-1.000] |
| Region 9 | .995 [.987-.999] | .997 [.990-.999] | .997 [.992-.999] |
| Region 10 | .998 [.995-1.000] | .998 [.995-1.000] | .994 [.983-.999] |

*Note.* Values in brackets represent the 95% confidence interval

**Supplementary Table 6.** Inter-Scanner Reliability Represented by Intraclass Correlation Coefficients - Skin Type I-II subjects (N=8)

|  | Full Data |  | No Vitiligo area =0 measurements |
| --- | --- | --- | --- |
|  | Scanners 1-3 | Scanners 2-4 | Scanners 1-3 |
| Region 1 | .992 [.972-.998] | ** | .977 [.873-.998] |
| Region 2 | .979 [.836-.997] | ** | .979 [.836-.997] |
| Region 3 | .997 [.989-.999] | ** | .996 [.983-.999] |
| Region 4 | .974 [.912-.995] | ** | .968 [.865-.996] |
| Region 5 | * | ** | ** |
| Region 6 | .983 [.943-.997] | ** | .973 [.727-1.000] |
| Region 7 | * | ** | ** |
| Region 8 | .963 [.878-.993] | ** | ** |
| Region 9 | .995 [.980-.999] | ** | .993 [.968-.999] |
| Region 10 | .999 [.995-1.000] | ** | .991 [.955-.999] |

Note. Values in brackets represent the 95% confidence interval; \*Correlation coefficient was not computed due to zero variance items; \*\*Too few cases for analysis (N<2)

**Supplementary Table 7.**
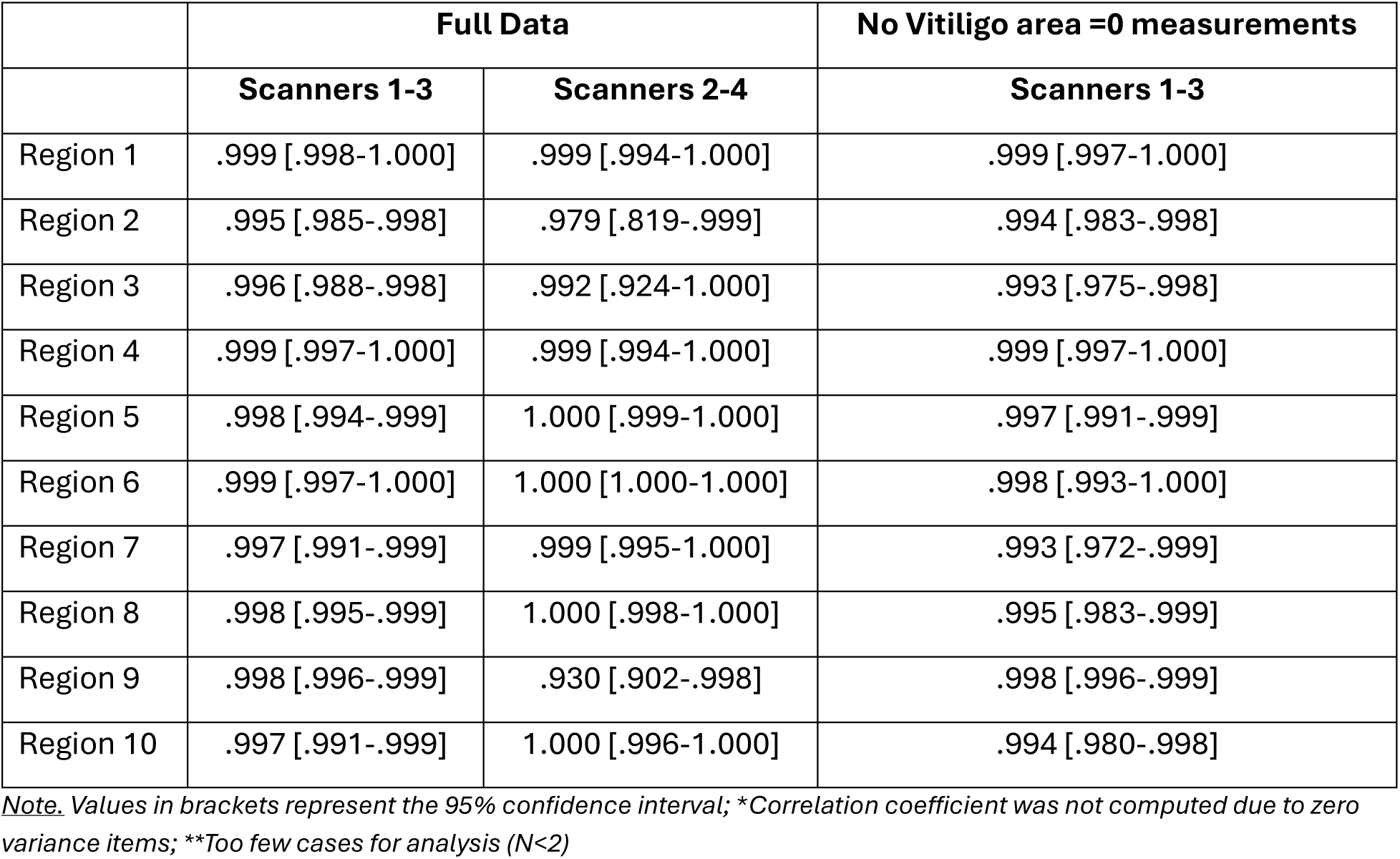
Inter-Scanner Reliability Represented by Intraclass Correlation Coefficients - Skin Type III-IV subjects (N=17)

|  | Full Data |  | No Vitiligo area =0 measurements |
| --- | --- | --- | --- |
|  | Scanners 1-3 | Scanners 2-4 | Scanners 1-3 |
| Region 1 | .999 [.998-1.000] | .999 [.994-1.000] | .999 [.997-1.000] |
| Region 2 | .995 [.985-.998] | .979 [.819-.999] | .994 [.983-.998] |
| Region 3 | .996 [.988-.998] | .992 [.924-1.000] | .993 [.975-.998] |
| Region 4 | .999 [.997-1.000] | .999 [.994-1.000] | .999 [.997-1.000] |
| Region 5 | .998 [.994-.999] | 1.000 [.999-1.000] | .997 [.991-.999] |
| Region 6 | .999 [.997-1.000] | 1.000 [1.000-1.000] | .998 [.993-1.000] |
| Region 7 | .997 [.991-.999] | .999 [.995-1.000] | .993 [.972-.999] |
| Region 8 | .998 [.995-.999] | 1.000 [.998-1.000] | .995 [.983-.999] |
| Region 9 | .998 [.996-.999] | .930 [.902-.998] | .998 [.996-.999] |
| Region 10 | .997 [.991-.999] | 1.000 [.996-1.000] | .994 [.980-.998] |

*Note.* Values in brackets represent the 95% confidence interval; \*Correlation coefficient was not computed due to zero variance items; \*\*Too few cases for analysis (N<2)

**Supplementary Table 8.** Intra-Scanner Reliability Represented by Intraclass Correlation Coefficients - Skin Type I-II subjects (N=8), no Vitiligo area = 0 Measurements.

|  | Scanner 1 | Scanner 2 | Scanner 3 |
| --- | --- | --- | --- |
| Region 1 | .985 [.922-1.000] | .992 [.955-1.000] | .984 [.935-.999] |
| Region 2 | .976 [.920-.997] | .978 [.932-.996] | .987 [.962-.997] |
| Region 3 | .978 [.915-.998] | .986 [.952-.998] | .992 [.973-.999] |
| Region 4 | .997 [.980-1.000] | .995 [.975-1.000] | .990 [.965-.999] |
| Region 5 | ** | ** | ** |
| Region 6 | .952 [.698-1.000] | .922 [.683-.998] | .872 [.542-.996] |
| Region 7 | ** | ** | ** |
| Region 8 | ** | .987 [.903-1.000] | .975 [.828-1.000] |
| Region 9 | .971 [.887-.998] | .968 [.885-.996] | .975 [.925-.996] |
| Region 10 | .990 [.937-1.000] | .986 [.943-.999] | .989 [.962-.999] |

*Note.* Values in brackets represent the 95% confidence interval; \*\*Too few cases for analysis (N<2)

**Supplementary Table 9.** Intra-Scanner Reliability Represented by Intraclass Correlation Coefficients – Skin Type III-IV subjects (N=17), no Vitiligo area = 0 Measurements.

|  | Scanner 1 | Scanner 2 | Scanner 3 |
| --- | --- | --- | --- |
| Region 1 | .977 [.930-.996] | .976 [.928-.996] | .985 [.954-.998] |
| Region 2 | .993 [.980-.999] | .997 [.990-.999] | .995 [.985-.999] |
| Region 3 | .993 [.972-1.000] | .998 [.990-1.000] | .997 [.987-1.000] |
| Region 4 | .996 [.989-.999] | .997 [.991-.999] | .998 [.993-1.000] |
| Region 5 | .989 [.955-.999] | .981 [.926-.999] | .967 [.849-.999] |
| Region 6 | ** | .908 [.358-1.000] | .950 [.700-1.000] |
| Region 7 | .995 [.965-1.000] | .998 [.983-1.000] | .995 [.959-1.000] |
| Region 8 | .990 [.921-1.000] | .998 [.985-1.000] | .992 [.925-1.000] |
| Region 9 | .994 [.984-.999] | .996 [.988-.999] | .997 [.991-1.000] |
| Region 10 | .996 [.986-1.000] | .996 [.986-1.000] | .988 [.953-.999] |

*Note.* Values in brackets represent the 95% confidence interval; \*\*Too few cases for analysis ( $N < 2$ )

